# Training residents in underwater colonoscopies is more effective than in air colonoscopies

**DOI:** 10.1101/2020.06.06.20124339

**Authors:** Dinesh Vyas, Josiah Chang, Megha Goyal, Nashwan Obad, Prakash Ramdass, Soujanya Sodavarapu

## Abstract

**Background:** Colonoscopy screenings are the most valuable tool in preventing colorectal mortalities. The traditional technique uses air-insufflation, but water-infusion is a newer colonoscopy technique which is rapidly becoming standard of care, as it may decrease patient discomfort and the need for analgesics and anesthetics. Research is still ongoing as to the comparability of detection rates between the two techniques. The purpose of this study was to determine if training residents in underwater colonoscopies is more effective than training them in traditional air-insufflation colonoscopies.

**Methods:** This study was a retrospective, single-institution study that compared the patient-related and procedure-related variables of 183 colonoscopies performed by two cohorts of physicians. In the first cohort, the gastroenterologist with a resident trainee performed an air colonoscopy. In the second, the gastroenterologist and resident trainee performed an underwater colonoscopy.

**Results:** For patient-related variables, there was no significant difference in age, previous abdominal surgeries, or bowel preparation. There were more females in the underwater group, which is significant as females tend to be harder to scope due to the increased tortuosity of their colon. For procedural outcomes, there was no significant difference in adenoma detection rate, cecal intubation rate, or procedural complications (hypotension, bradycardia). On average, the water colonoscopies required less midazolam and fentanyl, although they did have a longer procedural time.

**Conclusions:** Overall, these findings suggest that training residents in underwater colonoscopies may increase patient comfort and decrease complications with comparable success rates.

## Background

With over 50,000 colorectal cancer deaths per year in the US, colonoscopy screenings are the most valuable tool in detecting and preventing colorectal mortalities (1). As such, over 15 million colonoscopies are performed in the United States annually (2). The traditional procedure is an air insufflation colonoscopy, which has its issues regarding patient comfort and ease of approach. Air is used to inflate the colon, allowing for better visualization of the lumen as well as enabling the advancement of the scope. However, this technique can cause significant discomfort in patients, and thus generally requires sedation and analgesics to be administered to the patient. This poses its own problem in that it may mask the pain and discomfort which can be a telling sign of perforation (3). Another common complication of air insufflation colonoscopy is the risk of looping, in which the volume of air used to distend the colon can cause the colonoscope, along with the colon, to form a loop. This causes significant patient discomfort while also increasing the cost and overall procedure time. Lastly, not all of the air can be removed at the completion of the surgery, causing post-procedure discomfort to the patient in the form of cramps and abdominal bloating.

**Figure 1A.**
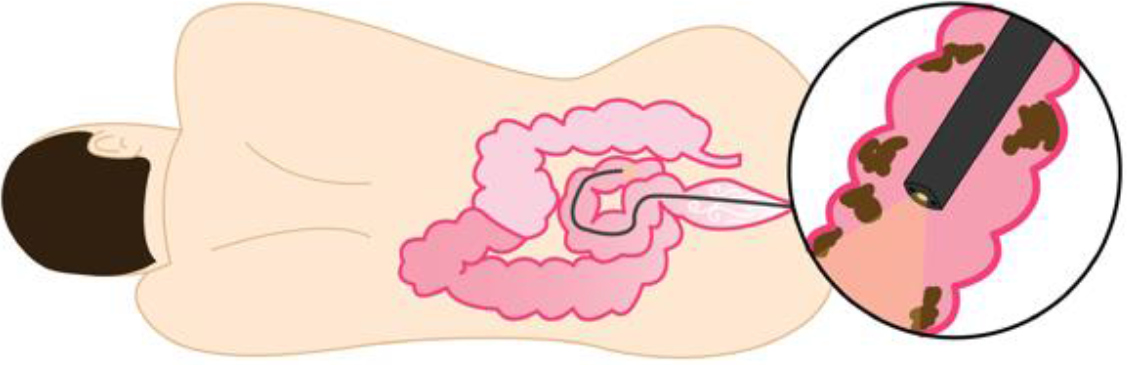
Air-insufflation colonoscopy.

**Figure 1B.**
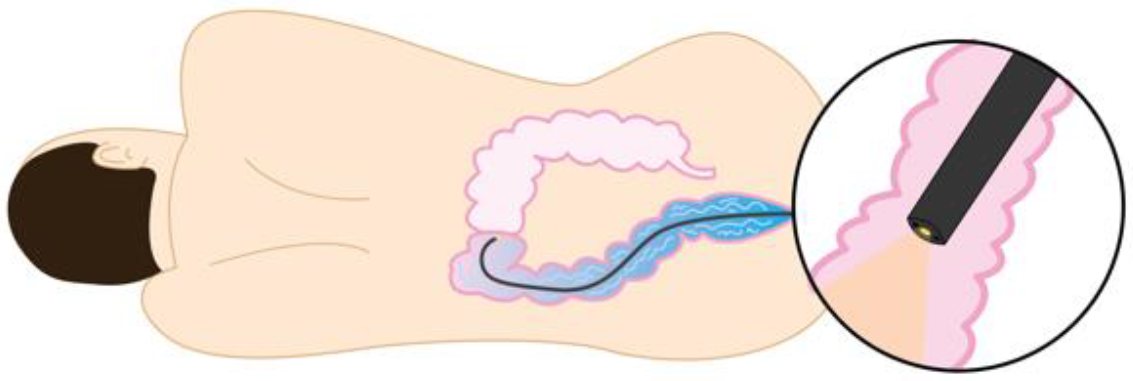
Water-infution colonoscopy.

All these potential issues can be daunting to the patient and likely contributes to the high number of patients who do not undergo a colonoscopy despite being indicated for one. It is estimated that only half of all patients referred for a colonoscopy follow through and complete the procedure, with one of the major reported barriers being fear of pain and discomfort (4). By decreasing these parameters, underwater colonoscopies have the potential to increase patient adherence to colonoscopy referrals.

In an underwater colonoscopy, water is used in the place of air to distend the colon. Firstly, this allows for better visualization of the colon, as water can also be used to flush out and clean the lumen of the colon. Water-infusion colonoscopies also greatly decrease the risk of looping, as gravity weighs the water-filled colon down and straightens out the colon. This, along with the natural lubricating advantage water-infusion holds over air-insufflation, means that patients would be less likely to experience pain and discomfort, and thus would require less sedation and analgesia (5). This in turn lowers the cost of the procedure and decreases the chance for significant complications such as bleeding and perforation. Post-procedurally, there are also less issues for patients undergoing water-infusion colonoscopy, as they are less likely to experience the cramping and bloating that an air-insufflation colonoscopy might cause. Furthermore, due to the increased ease of insertion and decreased risk of complications such as looping or perforation, training endoscopists in water-infusion colonoscopies may be easier and more efficacious than training them in air-insufflation colonoscopies in terms of resources, training time required, and procedure efficiency.

In total, underwater colonoscopies are a technique which may be easier to teach while also decreasing patient discomfort and potentially being more effective. There are several measures that can be used to determine the success of a colonoscopy, but two of the major ones that are commonly used are the adenoma detection rate (ADR) and the cecal intubation rate. The adenoma detection rate is the proportion of patients at or over the age of 50 who have a screening colonoscopy in which at least one precancerous polyp is detected. As the detection and prevention of colorectal cancer is the ultimate goal of colonoscopy screening, ADR is a useful measure of their success rate, and is recommended to be at least 20%(6). The cecal intubation rate is the proportion of colonoscopies in which the tip of the colonoscope passes proximally to the ileocecal valve. It is useful as an indicator of a complete examination of the lumen of the colon, and has a recommended benchmark of 90%for all colonoscopies, increased to 95%for screening colonoscopies (6). The aim of this study was twofold: (1) to determine if training residents in water-infusion versus air-insufflation would lead to a difference in patient preparation and outcome, particularly as measured by ADR and cecal intubation rate, and (2), if there was a difference in procedure time between the two groups.

## Methods

### Study population

This study was a retrospective, single-institution study that compared patient-related and procedure-related variables of a total of 183 patients within 2 cohorts of physicians. Of these patients, 100 had an air-insufflation colonoscopy performed by a gastroenterologist along with a resident (GE + R [air]), and 83 had a water-infusion colonoscopy performed by a gastroenterologist along with a resident (GE + R [water]).

The patient-related variables that were included were age, sex, presence of comorbidities, history of heart disease, lung disease, and kidney disease, and past history of abdominal surgery. The recorded procedure-related variables were level of bowel preparation, use of sedatives (propofol), use of analgesics (midazolam and fentanyl), hypotension, bradycardia, major/minor complications, incidence of diverticula, adenomatous polyps detected, cecal intubation rate, and time of procedure.

### Surgical methods

As seen in chart 2, the two groups differed slightly in procedure, with the majority of the GE + R (air) colonoscopies being finished by the gastroenterologist, while the majority of the GE + R (water) colonoscopies were finished by the resident.

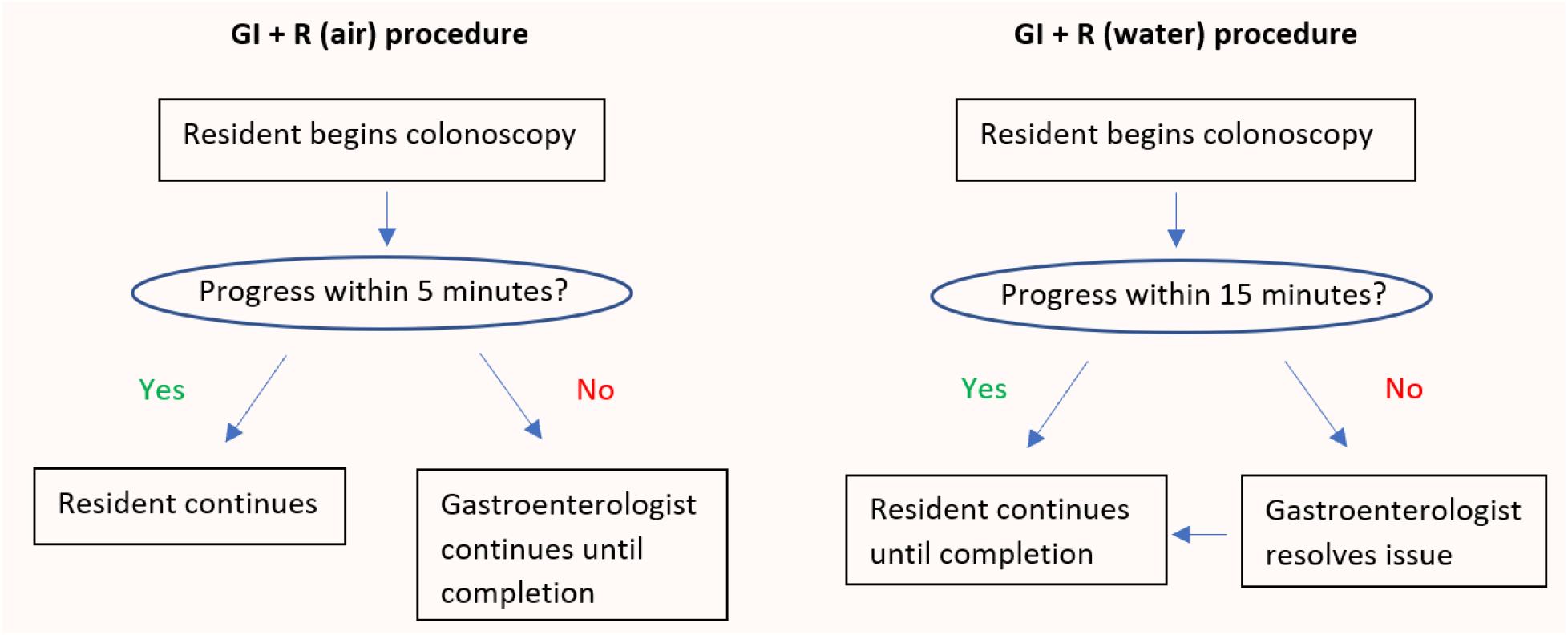

### Statistical analysis

Descriptive statistics (mean ± standard deviation [SD] or percentages) were computed for socio-demographic variables and clinical characteristics among the groups of physicians who performed the colonoscopy procedure. One-way ANOVA was used to compare continuous variables and the Χ2 (Cochran–Mantel–Haenszel) test was used to compare proportions between groups. All variables were reported as statistically significant at the 0.05 significance level. Analyses were performed using SPSS version 24 (IBM Corp., Armonk, NY, USA).

## Results

The mean ages and BMI were comparable between the two experimental groups. The underwater resident group had a greater proportion of females (47%, *n* = 39) compared to the air resident group (24%, *n* = 24, *p* = 0.001). There was also a significant difference in the indication for procedure between the two groups (Table 1, *p* = 0.002), as a larger proportion of the GE + R (water) group was undergoing screening compared to the GE + R (air) group, which had a greater proportion of patients indicated for procedure due to rectal bleeding. The two groups did have similar rates of previous endoscopic procedure as well as previous abdominal surgeries. There was not a significant difference in the bowel preparation between the two groups (*p* = 0.301).

**Table 1.** Patient-related variables, comparison between underwater and air colonoscopy

| Variables | GE + R (Water) | GE + R (Air) | p Values |
| --- | --- | --- | --- |
| Age, years, mean (SD) | 59.9 (9.9) | 57.3 (9.8) | 0.073 |
| Age, years, n (%) |  |  | 0.100 |
| ≤ 50 | 8 (9.6) | 21 (21.0) |  |
| 51 – 60 | 29 (34.9) | 39 (39.0) |  |
| 61 – 70 | 38 (45.8) | 34 (34.0) |  |
| > 70 | 8 (9.6) | 6 (6.0) |  |
| Gender, n (%) |  |  | **0.001** |
| Female | 39 (47.0) | 24 (24.0) |  |
| Male | 44 (53.0) | 76 (76.0) |  |
| Race, n (%) |  |  | **0.011** |
| Hispanic | 30 (36.1) | 42 (42.0) |  |
| Caucasian | 18 (21.7) | 18 (18.0) |  |
| African American | 12 (14.5) | 21 (21.0) |  |
| Asian | 21 (25.3) | 9 (9.0) |  |
| Other | 2 (2.4) | 10 (10.0) |  |
| Indication, n (%) |  |  | **0.002** |
| Screening | 62 (74.7) | 51 (51.0) |  |
| Rectal Bleeding | 7 (8.4) | 24 (24.0) |  |
| Other | 14 (16.9) | 25 (25.0) |  |
| Previous Scope, n (%) |  |  | 0.278 |
| Yes | 13 (15.7) | 22 (22.0) |  |
| No | 70 (84.3) | 78 (78.0) |  |
| Bowel Preparation, n (%) |  |  | 0.301 |
| Good | 58 (69.9) | 61 (61.0) |  |
| Fair | 8 (9.6) | 17 (17.0) |  |
| Poor | 17 (20.5) | 22 (22.0) |  |
| Previous Abdominal Surgery, n (%) |  |  | 0.328 |
| Yes | 25 (30.1) | 37 (37.0) |  |
| No | 58 (69.9) | 63 (63.0) |  |
| Hemorrhoids |  |  | **<0.001** |
| Yes | 33 (39.8) | 87 (87.0) |  |
| No | 50 (60.2) | 13 (13.0) |  |
| Comorbidities, n (%) |  |  | **0.017** |
| None | 8 (9.6) | 1 (1.0) |  |
| 1-2 | 34 (41.0) | 53 (53.0) |  |
| ≥ 3 | 41 (49.4) | 46 (46.0) |  |
| Body mass index, kg/m<sup>2</sup>, (SD) | 29.9 (6.7) | 31.1 (6.4) | 0.203 |

*p* < 0.05, significant values in bold

As for measurements of the procedure success rate between the two resident groups, the underwater group had comparable polyp detection rates (48%) to the air group, which had a 54%polyp detection rate (*p* = 0.603). The cecal intubation rate was also similar between the two groups, with the water group at 94%and the air group at 96%(*p* = 0.384). The GE + R (water) colonoscopies did have a higher mean procedural time compared to the GE + R (air) colonoscopies, at 35.7 minutes and 27.4 minutes respectively (p < 0.001). There was no significant difference in the rate of diverticulosis (*p* = 0.144), but there was a much higher rate of hemorrhoids in the GE + R (air) group (87%) than in the GE + R (water) group (40%, *p* < 0.001). Both groups showed similar rates of hypotension (Water group = 49%, Air group = 38%; *p* = 0.121) and bradycardia (Water group = 30%, Air group = 34%, *p* = 0.576) throughout the procedure. Lastly, the amount of propofol administered was similar in the two groups, but less midazolam and fentanyl was required in the GE + R (water) group as compared to the air group (Table 2, *p* < 0.001 for midazolam, *p* < 0.001 for fentanyl).

**Table 2.** Procedure-related variables, comparison between underwater and air colonoscopy

| Variables | GE + R (Water) | GE + R (Air) | p Values |
| --- | --- | --- | --- |
| Cecal Intubation |  |  | 0.384 |
| Yes | 78 (94.0) | 96 (96.0) |  |
| No | 5 (6.0) | 4 (4.0) |  |
| Diverticulosis |  |  | 0.144 |
| Yes | 20 (24.1) | 34 (34.0) |  |
| No | 63 (75.9) | 66 (66.0) |  |
| Hemorrhoids |  |  | **<0.001** |
| Yes | 33 (39.8) | 87 (87.0) |  |
| No | 50 (60.2) | 13 (13.0) |  |
| Hypotension |  |  | 0.121 |
| Yes | 41 (49.4) | 38 (38.0) |  |
| No | 42 (50.6) | 62 (62.0) |  |
| Bradycardia |  |  | 0.576 |
| Yes | 25 (30.1) | 34 (34.0) |  |
| No | 58 (69.9) | 66 (66.0) |  |
| Number of Adenomatous Polyps |  |  | 0.603 |
| None | 43 (51.8) | 46 (46.0) |  |
| 1-2 | 27 (32.5) | 33 (33.0) |  |
| ≥ 3 | 13 (15.7) | 21 (21.0) |  |
| Number of polyps, mean, (SD) | 1.1 (1.7) | 1.7 (2.9) | 0.080 |
| Total Time, mins, (SD) | 35.7 (14.9) | 27.4 (12.3) | **<0.001** |
| Propofol, mg, (SD) | 363.5 (225.8) | 346.1 (185.9) | 0.651 |
| Midazolam, mg, (SD) | 2.0 (0.0) | 5.8 (2.5) | **<0.001** |
| Fentanyl, mcg, (SD) | 95.3 (13.6) | 134.7 (91.3) | **0.003** |

*p* < 0.05, significant values in bold

## Discussion

Traditional air insufflation colonoscopies have been the classic technique for endoscopies, but recent studies suggest that water infusion colonoscopies are more effective in reducing patient pain as well as improve colonoscopy success rates. In a randomized controlled trial involving 230 patients, Radaelli et. al [7] reported that water-infusion patients required less on-demand sedation with lower pain scores than air-insufflation patients. In addition to supporting these results, a randomized controlled trial by Cadoni et. al [8] involving 672 patients also reported higher overall adenoma detection rates and similar cecal intubation rates.

There are also several other factors that must be taken into account when trying to determine colonoscopy success rates. In a retrospective review of 5125 colonoscopies between 1979-1995, Dafnis et. al [9] reported female sex, increased age, and presence of diverticulosis as factors negatively influencing completion rate, while patients with previous colonic surgeries positively affected the completion rate. In a more recent review of 5352 patients, Hsu et. al [9] reported patient age > 60 years as a negative factor for cecal intubation rate and quality of bowel prep as a positive factor, while patient sex, BMI, and prior abdominal surgeries did not appear to have a significant effect on cecal intubation rate.

While it was hypothesized that the GE + R (water) group might have better bowel preparation than the other two groups due to the ability to use water to clean the bowel, there was no significant difference in bowel preparation in this present study. Females are generally regarded as more difficult to scope due to the tortuosity of the female colon – however, despite having a higher proportion of female patients in the GE + R (water) group, the success rate as measured in both adenoma detection rate and cecal intubation rate was comparable to the GE + R (air) group. It should be noted that the GE + R (water) group did have a longer procedural time than the GE + R (air group).

## Conclusion

When performing water infusion colonoscopies, residents training with a gastroenterologist had similar adenoma detection rates and cecal intubation rates as when performing air insufflation colonoscopies. In total, the findings of this present study suggest that training residents in underwater colonoscopies may be more efficacious in increasing patient comfort and decreasing procedural complications without sacrificing success rate, but with a slight increase in procedure time.

However, given the limited sample size, the variation in patient populations between the cohorts, and its retrospective nature, the results of this study are far from conclusive. Further research with randomized controlled studies involving larger patient populations are necessary to draw more definite conclusions about the efficacy of training residents in underwater colonoscopies as compared to the traditional air insufflation colonoscopies.

## Data Availability

The authors confirm that the data supporting the findings of this study are available within the article.

